# FAIR-IFICATION OF STRUCTURED CLINICAL DATA

**DOI:** 10.1101/2021.07.23.21261032

**Authors:** Varsha Gouthamchand, Andre Dekker, Leonard Wee, Johan van Soest

**Affiliations:** Radiotherapy (MAASTRO), School of Oncology, Maastricht University Medical Centre+, Maastricht, The Netherlands; Clinical Data Science, Maastricht University, Maastricht, The Netherlands

**Keywords:** FAIR, Knowledge graphs, Linked Data, Semantic Web, SPARQL, RDF triples, Ontologies

## Abstract

One of the common concerns in clinical research is improving the infrastructure to facilitate the reuse of clinical data and deal with interoperability issues. FAIR (Findable, Accessible, Interoperable and Reusable) Data Principles enables reuse of data by providing us with descriptive metadata, explaining what the data represents and where the data can be found. In addition to aiding scholars, FAIR guidelines also help in enhancing the machine-readability of data, making it easier for machine algorithms to find and utilize the data. Hence, the feasibility of accurate interpretation of data is higher and this helps in obtaining maximum results from research work. FAIR-ification is done by embedding knowledge on data. This could be achieved by annotating the data using terminologies and concepts from Web Ontology Language (OWL). By attaching a terminological value, we add semantics to a specific data element, increasing the interoperability and reuse. However, this FAIR-ification of data can be a complicated and a time-consuming process. Our main objective is to disentangle the process of making data FAIR by using both domain and technical expertise. We apply this process in a workflow which involves FAIR-ification of four independent public HNSCC datasets from The Cancer Imaging Archive (TCIA). This approach converts the data from the four datasets into Linked Data using RDF triples, and finally annotates these datasets using standardized terminologies. By annotating them, we link all the four datasets together using their semantics and thus a single query would get the intended information from all the datasets.

## Introduction

The world is at the realm of secondary use of real-world data. With the rapid growth of healthcare data available across the world [1], there is an enormous potential for the “secondary use or reuse” of this data as this leads to a more time-effective and cost-effective research. Collected during clinical trials or as healthcare records, the secondary usage of this data is generally for purposes other than its original use [2]. The potential use cases involve using this data for enhancing the delivery of health care for individuals and broaden the existing knowledge about diseases and their respective treatments, by increasing research and analysis of such data [3]. Reuse of clinical data has been prevalent for decades and has proven effective in detecting early stages of a disease, predicting patient’s length of stay or patient’s overall survival [4]. Despite the massive amount of clinical data generated every year, the secondary usage of these data is very limited.

Moreover, when patients with rare diseases are considered, they are often at a disadvantage due to the rarity of the disease. Rare diseases bring about lesser data with very limited clinical research and management [5]. According to [6], it was found that the five-year relative survival rate of patients with rare cancer was 48.5%, whereas for patients with other common cancers was 63.4%. In addition to this, the volume of publicly “donated data” (e.g., in TCIA or TCGA) is very small relative to total amount of clinical data generated in CRFs. A “big dataset” in rare cancers consists of data from a few hundred patients, which is a separate scale from social media, twitter text or natural photographs numbering in the many millions. Hence, data quality and semantics play a larger role in healthcare. Moreover, this small amount of data contains personal/idiosyncratic coding and schema, and data being structured already assumes a very high degree of implicit knowledge. Therefore, how do we use and share this data, in a privacy-preserving way, without doctors/investigators losing control of their own data? How do we make this a community effort among different domains of expert knowledge? (i.e., Clinical versus Machine Learning versus Data Science).

FAIR (Findable, Accessible, Interoperable, and Reusable) Data Principles were designed to support reusability of scholarly data. FAIR provides an infrastructure, which enables maximum usefulness of research data, by providing descriptive metadata about the data. This helps in structuring the information in the data in a way that it is interpretable by machine algorithms, without direct human supervision [7]. It is to be noted that FAIR does not necessarily imply Open-ness of data, with the key difference between both FAIR and Open data being the term “access”. According to FAIR, data is accessible only for a defined set of people, for example a group of researchers and clinicians working together [8]. Whereas according to Open Data Handbook, “*Open data is data that can be freely used, re-used and redistributed by anyone - subject only, at most, to the requirement to attribute and sharealike”* [9]. This also applies for the opposite case, implying that Open Data is not always FAIR.

FAIR data deals with the challenge of enabling syntactic and semantic interoperability for comprehensive and reusable processing of big data. Syntactic interoperability refers to a standardized communication and data exchange between two or more systems (this could be done with different interfaces and programming languages). Semantic interoperability is, when the exchanged data is understood by the systems involved, thus making it a more challenging yet desirable concept [10]. One of the main requirements to efficiently extract valuable information from large amounts of multi-centered clinical data is supporting both types of interoperability [11].

This is achieved using the concept of Linked Data. Linked Data is a set of guidelines for creating machine-readable and interlinked data on the Web. This collection of Linked Data is what makes up the Semantic Web [12]. For constructing Linked Data, data is often represented in standard Linked Data format called the RDF (Resource Description Framework). The RDF is a standard for defining relationships between data objects and interconnecting them. Every resource is identified by a URI (Universal Resource Identifier), which makes the data ‘linkable’. RDF data is represented in triples with a Subject, a Predicate and an Object [13].

RDF data can be stored in an RDF graph database (also called as semantic graph database or triple store), which acts as a query endpoint to get access to the data efficiently. Accessing data from these endpoints requires technologies like SPARQL (SPARQL Protocol and RDF Query Language), which is a W3C-standardized semantic query language to extract valuable information from a set of triples [14].

## Methods

### Description of the datasets

The data used in this submission is from the following data collections available on The Cancer Imaging Archive (TCIA).

1. RADIOMICS-HN1 from Maastro in Maastricht. This data collection has clinical data and Computed Tomography (CT) from 137 patients with head and neck squamous cell carcinoma (HNSCC), treated with radiotherapy [16].
2. HNSCC from MD Anderson Cancer Center in Houston, which contains a cohort of clinical meta-data and contrast-enhanced computed tomography (CECT) scans for 495 oropharyngeal cancer (OPC) patients [17].
3. OPC Radiomics from Princess Margaret Cancer Centre in Toronto. The collection includes data from 606 non-metastatic p16-positive oropharyngeal cancer (OPC) patients, treated with radiotherapy or chemo-radiotherapy between 2005 and 2010 [18].
4. HEAD-NECK-PET-CT from Montreal, which includes FDG-PET/CT and radiotherapy planning CT imaging data of 298 patients with head-and-neck cancer (H&N). Pre-treatment FDG-PET/CT scans for the patients were taken between April 2006 and November 2014 [19].

### FAIR data dashboard (demo module)

#### Step 1

The first step in the demo module is converting the data in the four datasets into RDF triples. This data conversion is done using our in-house developed software called the Triplifier [15]. Triplifier accepts a relational database as an input and gives out two files. One is the OWL file, which has the structure (a data-specific ontology). The other is the Output file with the actual instance information, which is the data from the dataset converted into RDF triples (as shown in fig 3).

**Fig 1.**
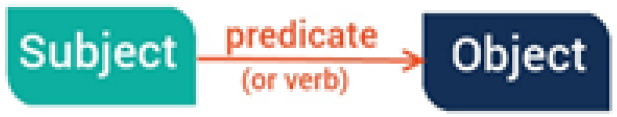
Format of an RDF triple

**Fig 2.**
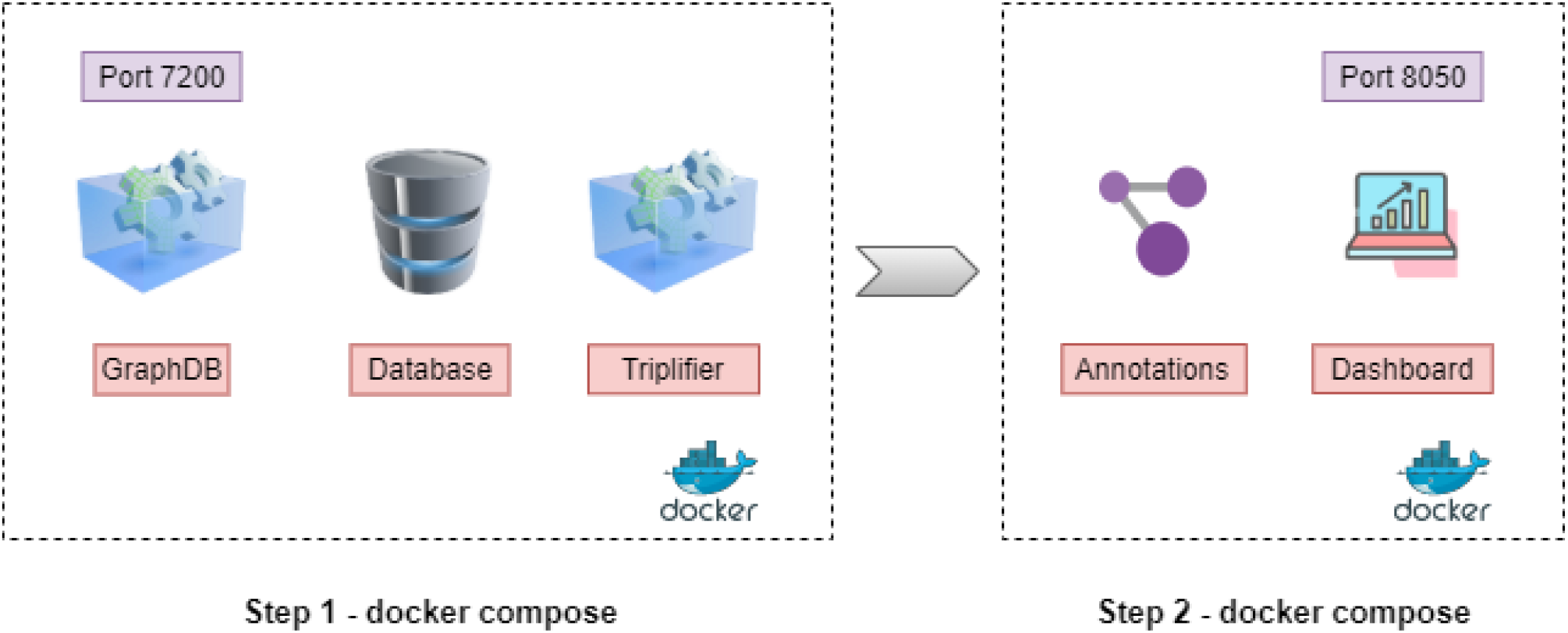
Workflow of FAIR data dashboard

**Fig 3.**
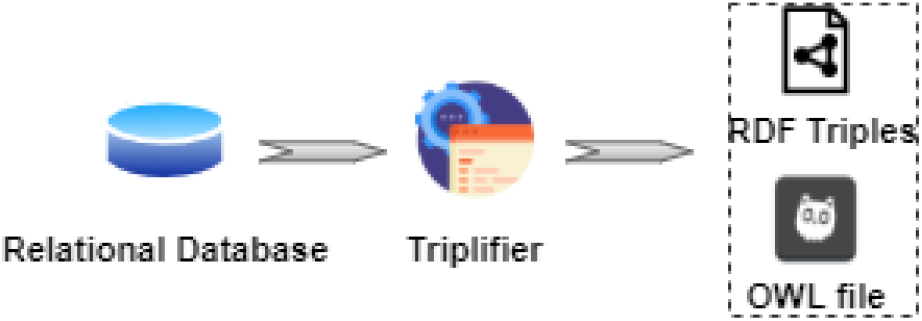
Workflow of Triplifier

In order to obtain the triples, the user can clone the code repository linked to this submission: https://gitlab.com/UM-CDS/projects/flyover_project/-/tree/MedRxiv_branch. In the command line interface, the user can navigate to the project folder and using the command ‘*docker-compose up –d’* will build four docker images – Postgres, pgAdmin, Triplifier and GraphDB. The docker files execute the following process:

- It builds a Postgres and a pgAdmin container.
- SQL insert queries immediately fire into Postgres and this creates four databases (can be checked in pgAdmin), one for each of the four datasets.
- Four parallel Triplifier projects fire up, one for each of the four clinical “projects”.
- Triplifier projects exit code upon successful completion.
- GraphDB (our local graph database) now has four RDF repositories, each with two graphs:
  – <http://data.local/> with the RDF triples.
  – <http://ontology.local/> with a data specific ontology.

#### Step 2

Upon the successfully completion of Triplifier, all the four graphDB repositories should consist of the RDF triples pertaining to each clinical project. Next, the second docker compose file can be built by navigating to the *annotations* folder inside the project https://gitlab.com/UM-CDS/projects/flyover_project/-/tree/MedRxiv_branch and the command ‘*docker-compose up –d’* would build the following two images:

- An annotations script that adds a new graph <http://annotation.local/> to all the four repositories in graphDB. This graph has triples with new semantic mappings for the flat RDF triples from the Triplifier.
- Finally, the user can visualize the data from all the four annotated datasets (now linked together via their semantics) in a dashboard running at port 8050 at their local system. This dashboard uses SPARQL queries to retrieve the triples from graphDB.

#### Construction of Annotations

The RDF triples from the Triplifier (fig 4) have a flat schema and does not hold any semantics to it. This is where ontologies come into play. Ontologies help in providing syntactic and semantic usability to the data. Ontologies can be viewed as a pre-defined data dictionary, which also contain information about how the entities in a field are related to each other. For our data, we use universal ontologies such as Radiation Oncology Ontology (ROO) and National Cancer Institute Thesaurus (NCIT). This process requires manual work and collaboration from the clinicians, language experts and the researchers as it is crucial to understand the clinical data (which could initially be in any language). The annotations include new meaningful predicates and class equivalencies from pre-defined ontologies which add semantics to the flat RDF triples from the Triplifier. This process is repeated for all the four datasets (refer fig 5-7).

**Fig 4.**
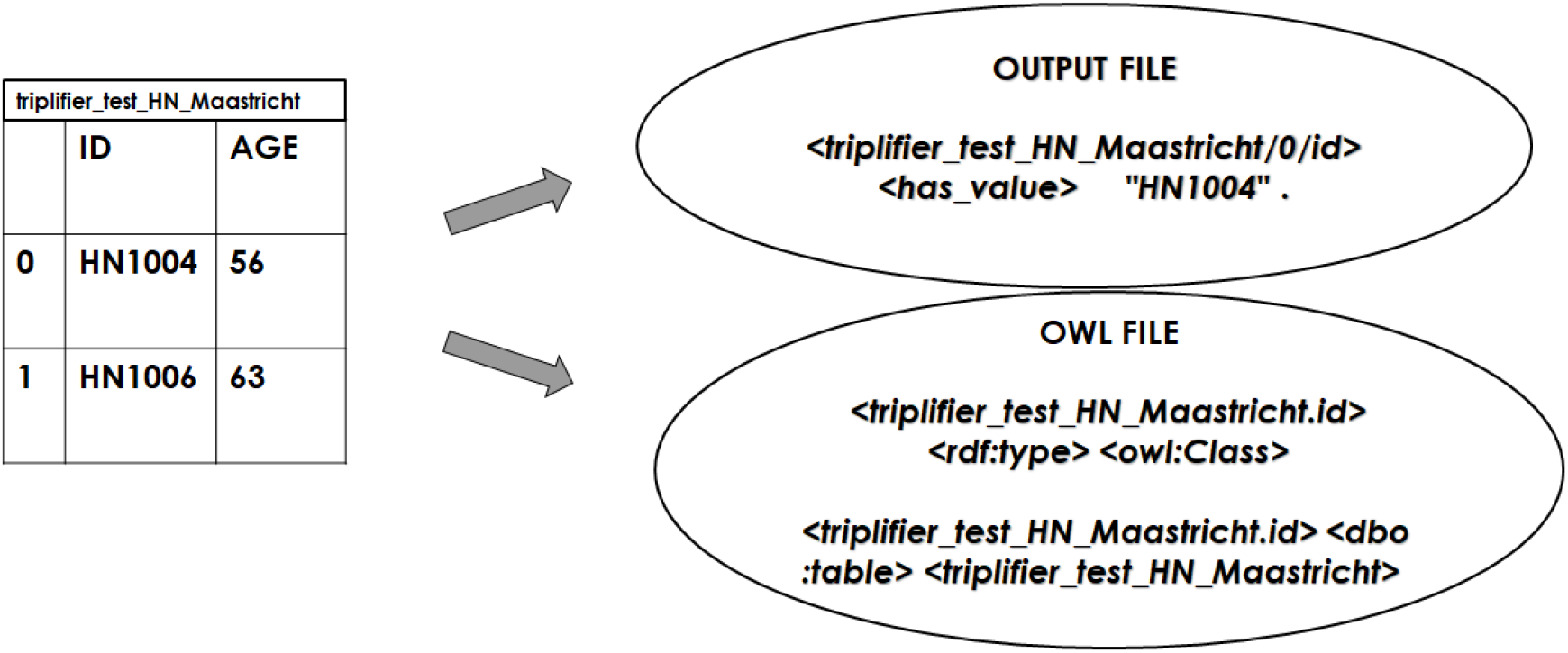
Output files format of Triplifier for HN_ONE (Maastro) dataset

**Fig 5.**
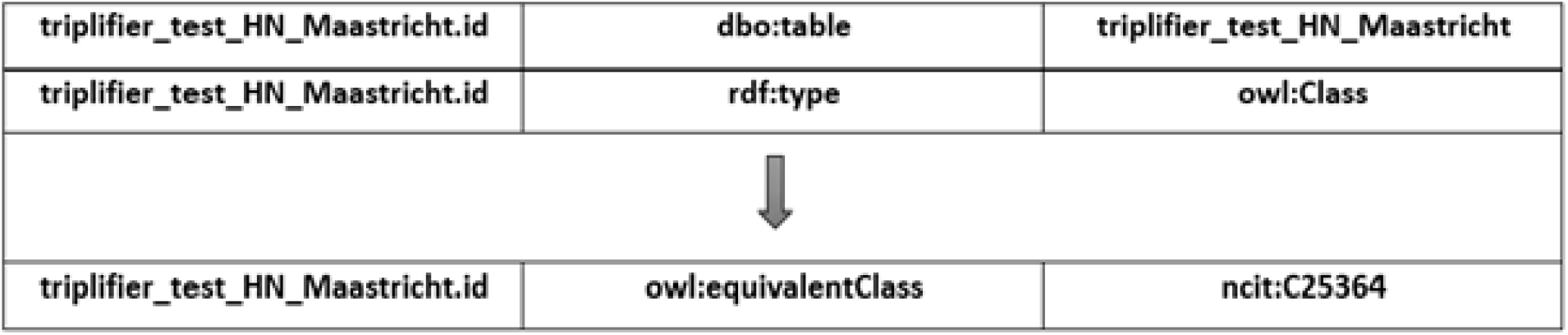
New class equivalencies from NCIT in annotations graph instead of the flat triples in Triplifier’s output. (C25364 – NCIT CODE FOR PATIENT IDENTIFIER)

**Fig 6.**
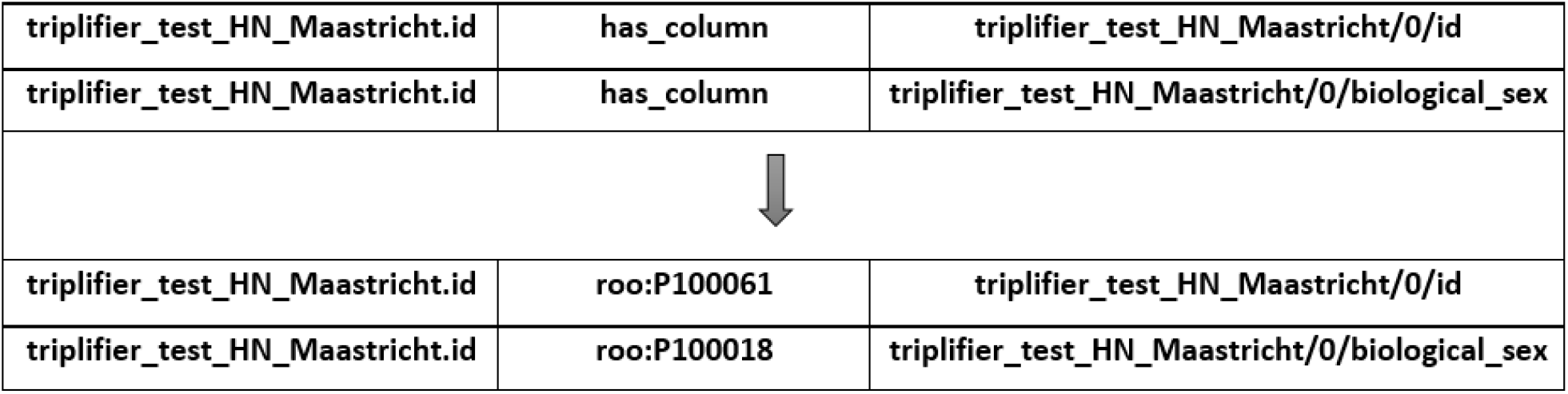
New meaningful predicates from ROO in annotations graph instead of the flat predicates in Triplifier’s output. (P100061 - ROO CODE FOR ‘HAS_IDENTIFIER’, P100018 – ROO CODE FOR ‘HAS_BIOLOGICAL_SEX’)

**Fig 7.**
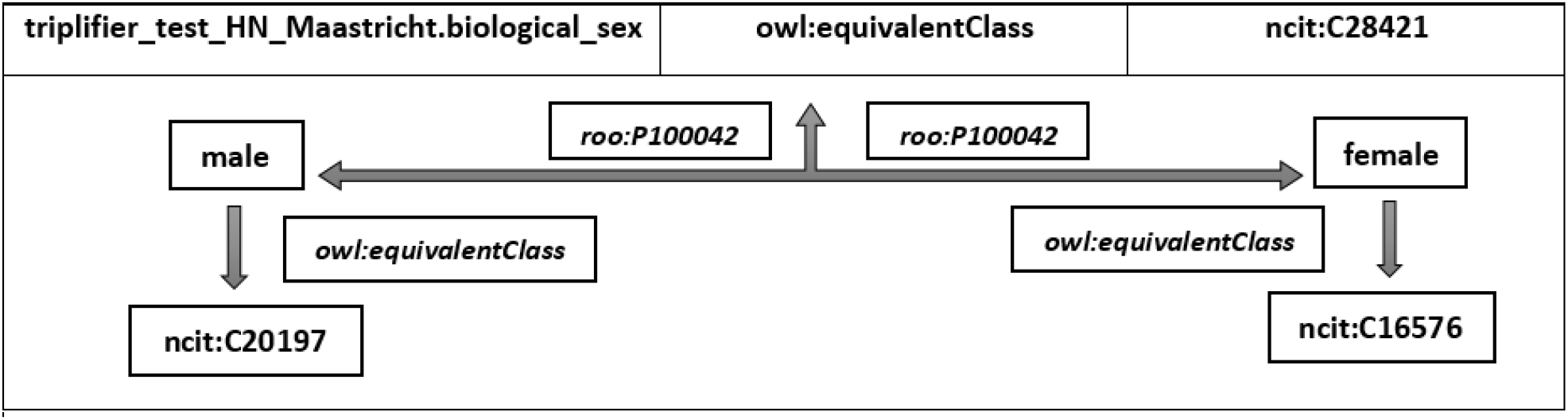
New class equivalencies for categorical columns. (P100042 – ROO CODE FOR ‘HAS_VALUE’, C28421 – NCIT CODE FOR BIOLOGICAL SEX, C20197 – NCIT CODE FOR MALE, C16576 – NCIT CODE FOR FEMALE)

### SPARQL endpoint

The annotations with the newly updated semantics and ontological tags for the RDF triples, are uploaded to the SPARQL endpoint GraphDB in their respective repositories (which already has the Output file and the OWL file). Now that meaningful annotations are added for all the four datasets and they are linked to their semantics, one common federated SPARQL query should be able to retrieve required results from all the datasets.

### Interactive Dashboard

As a potential use case of this project, we build an interactive visualization dashboard (as shown in fig 8) locally for the demo module, using the newly created Linked Data from all the four datasets, that lets the user choose to view the statistics of all the datasets (together or individually). This dashboard is created entirely using the results from federated SPARQL queries, which is then parsed through a Python program providing the clinicians with the output in the form of interactive graphs and charts.

**Fig 8.**
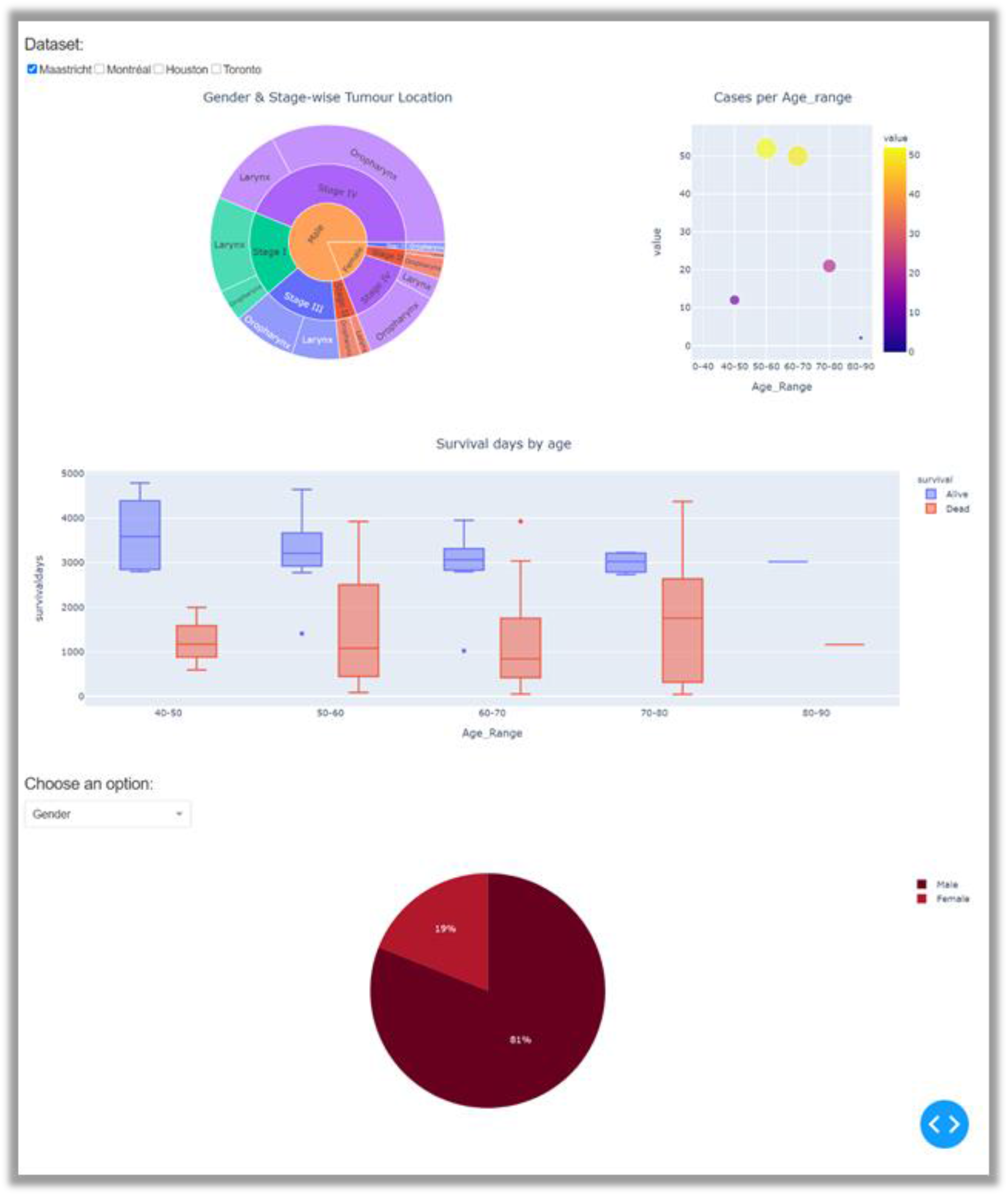
Visualization Dashboard

However, it is to be noted that this dashboard is only for DEMO purpose. It has been created locally by building a centralized data using the four repositories and it is not a privacy preserving distributed dashboard.

### Example SPARQL Queries

The first example we have considered is a researcher/user trying to get the ID values, gender, AJCC stage and the tumour location of patients from the FAIRified data. The following query would be able to get the mentioned values from the datasets after they have been semantically mapped with their ontologies. The user can copy and run the query in the SPARQL endpoint (GraphDB) running at their local host.

**Example 1.**
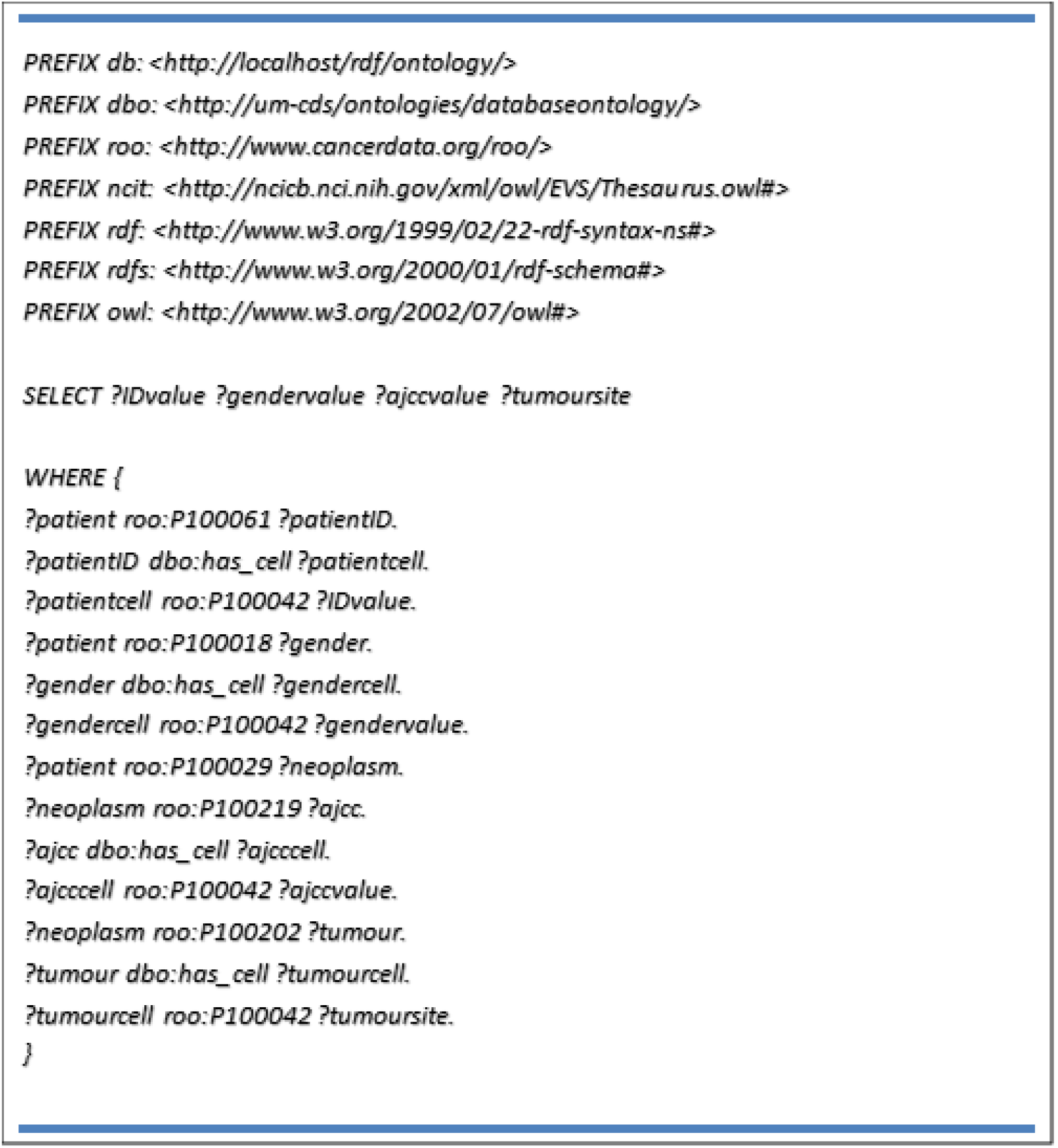
SPARQL query to get ID, gender, AJCC stage and Tumour Location

The second example we have considered is a researcher/user trying to get the values of T stage and M stage for individual patients from the annotated data. The following query would be able to get the mentioned values from the datasets after they have been semantically mapped with their ontologies.

**Example 2.**
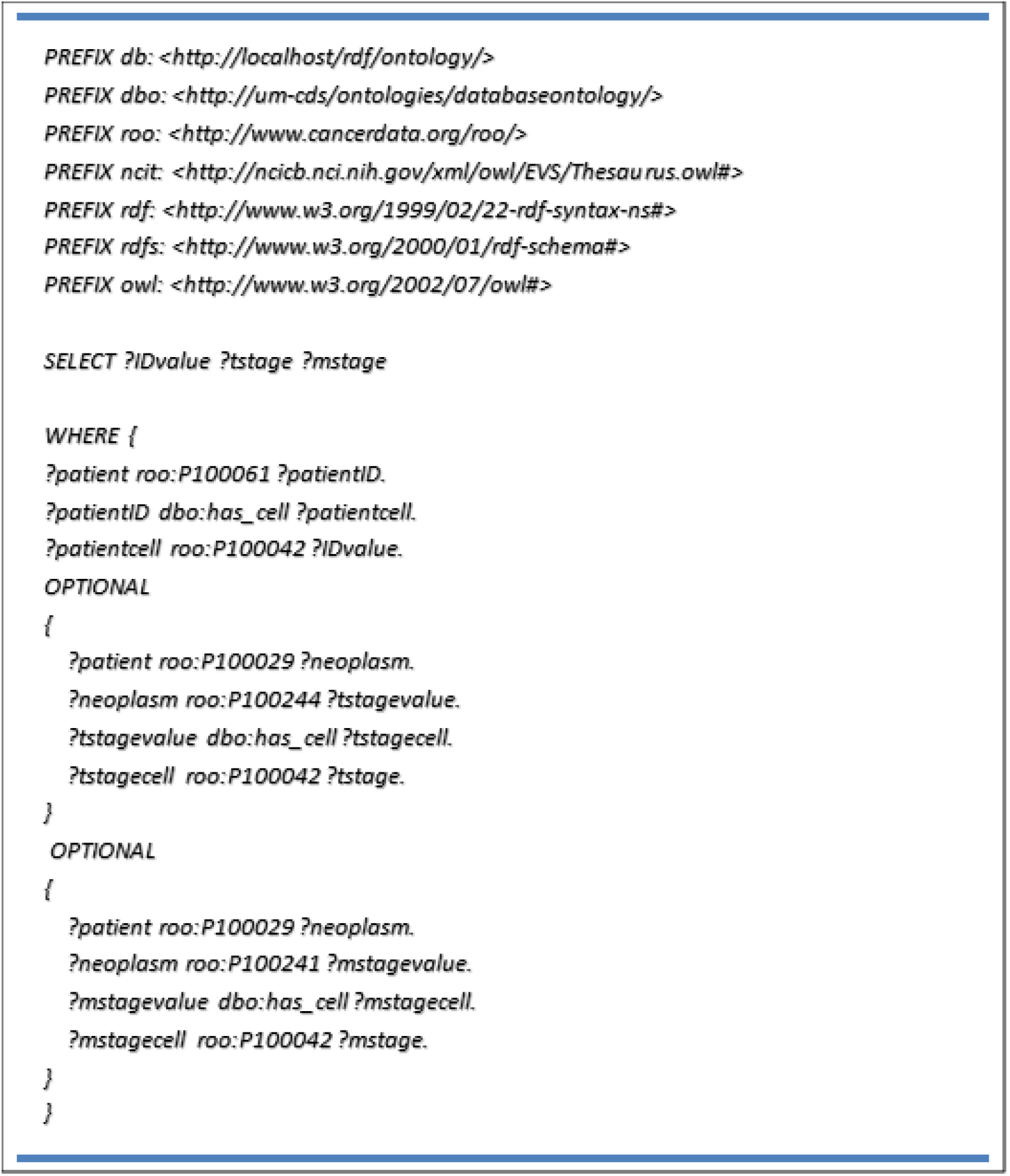
SPARQL query to get values of T-Stage and M-Stage for all patients

## Discussion and Conclusion

By designing the above process to create FAIR data from the datasets, we successfully achieve both syntactic interoperability and semantic interoperability in two manageable pieces. One by creating the standard RDF triples using the Triplifier and the other by creating an annotations graph with semantics for the triples.

The task requires cooperation from the user’s side for answering a few questions about the data and for gaining insights from the metadata for the FAIR-ification, but this does not demand any high degree of programming knowledge and tooling/technical skills from them, thus enabling an easy availability of FAIR data at the local stations.

## Data Availability

Data is already publicly available.

https://wiki.cancerimagingarchive.net/display/Public/HNSCC

https://wiki.cancerimagingarchive.net/pages/viewpage.action?pageId=33948764

https://wiki.cancerimagingarchive.net/display/Public/Head-Neck-Radiomics-HN1

https://wiki.cancerimagingarchive.net/display/Public/Head-Neck-PET-CT

## Notes

### Competing Interest Statement

The authors have declared no competing interest.

### Funding Statement

The work of author Varsha Gouthamchand has been supported financially by the Dutch Research Council (NWO) Indo-Dutch grant TRAIN (629.002.212).
The work of author Leonard Wee has been funded by the Hanarth Foundation.

### Author Declarations

Exemption - data is public open access and already freely available

### Summary of Updates

Updated the Funding Information.

## References

1) The healthcare data explosion. Accessed April 22, 2021. http://www.rbccm.com/en/gib/healthcare/story.page?dcr=templatedata/rbccm/episode/data/healthcare/the_healthcare_data_explosion.

2) A. Martani, L. D. Geneviève, C. Pauli-Magnus, S. McLennan, and B. S. Elger, “Regulating the Secondary Use of Data for Research: Arguments Against Genetic Exceptionalism,” Front. Genet., vol. 10, 2019, doi: 10.3389/fgene.2019.01254

3) C. Safran et al., “Toward a National Framework for the Secondary Use of Health Data: An American Medical Informatics Association White Paper,” J Am Med Inform Assoc, vol. 14, no. 1, pp. 1–9, 2007, doi: 10.1197/jamia.M2273

4) S. M. Meystre, C. Lovis, T. Bürkle, G. Tognola, A. Budrionis, and C. U. Lehmann, “Clinical Data Reuse or Secondary Use: Current Status and Potential Future Progress,” Yearb Med Inform, vol. 26, no. 1, pp. 38–52, Aug. 2017, doi: 10.15265/IY-2017-007

5) L. Botta et al., “Incidence and survival of rare cancers in the US and Europe,” Cancer Med, vol. 9, no. 15, pp. 5632–5642, May 2020, doi: 10.1002/cam4.3137

6) G. Gatta et al., “Burden and centralised treatment in Europe of rare tumours: results of RARECAREnet-a population-based study,” Lancet Oncol, vol. 18, no. 8, pp. 1022–1039, Aug. 2017, doi: 10.1016/S1470-2045(17)30445-X

7) M. D. Wilkinson et al., “The FAIR Guiding Principles for scientific data management and stewardship,” Scientific Data, vol. 3, no. 1, Art. no. 1, Mar. 2016, doi: 10.1038/sdata.2016.18

8) “Open Data and FAIR Data: differences and similarities | Plataforma OGoov,” OGoov Open Government Platform, May 23, 2019. https://www.ogoov.com/en/blog/open-data-and-fair-data-differences-and-similarities/ (accessed Apr. 27, 2021).

9) “What is Open Data?” http://opendatahandbook.org/guide/en/what-is-open-data/ (accessed Apr. 27, 2021).

10) “Syntactic and Semantic Interoperability | Electrosoft.” https://www.electrosoft-inc.com/resources/syntactic-and-semantic-interoperability (accessed Apr. 27, 2021).

11) M. Sudmanns, D. Tiede, S. Lang, and A. Baraldi, “Semantic and syntactic interoperability in online processing of big Earth observation data,” International Journal of Digital Earth, vol. 11, no. 1, pp. 95–112, Jan. 2018, doi: 10.1080/17538947.2017.1332112

12) “Data - W3C.” https://www.w3.org/standards/semanticweb/data (accessed Apr. 27, 2021).

13) “What is an RDF Triplestore? | Ontotext Fundamentals,” Ontotext. https://www.ontotext.com/knowledgehub/fundamentals/what-is-rdf-triplestore/.

14) “What Are Linked Data and Linked Open Data? | Ontotext Fundamentals,” Ontotext. https://www.ontotext.com/knowledgehub/fundamentals/linked-data-linked-open-data/ (accessed Apr. 27, 2021).

15) J. van Soest, A. Choudhury, N. Gaikwad, M. Sloep, and A. Dekker, “Annotation of existing databases using Semantic Web technologies: making data more FAIR,” p. 8.

16) Wee, Leonard, and Andre Dekker. “Data from Head-Neck-Radiomics-HN1.” The Cancer Imaging Archive, 2019. Accessed May 13, 2021. https://wiki.cancerimagingarchive.net/x/iBglAw.

17) Elhalawani, Hesham, Aubrey L. White, James Zafereo, Andrew J. Wong, Joel E. Berends, Shady AboHashem, Bowman Williams, et al. “Radiomics Outcome Prediction in Oropharyngeal Cancer.” The Cancer Imaging Archive, n.d. Accessed May 14, 2021. https://wiki.cancerimagingarchive.net/x/UAIGAg.

18) Kwan, Jennifer Yin Yee, Jie Su, Shao Hui Huang, Laleh S. Ghoraie, Wei Xu, Biu Chan, Kenneth W. Yip, et al. “Data from Radiomic Biomarkers to Refine Risk Models for Distant Metastasis in Oropharyngeal Carcinoma.” The Cancer Imaging Archive, 2019. Accessed May 14, 2021. https://wiki.cancerimagingarchive.net/x/XAQGAg.

19) Vallières, Martin, Emily Kay-Rivest, Léo Perrin, Xavier Liem, Christophe Furstoss, Nader Khaouam, Phuc Nguyen-Tan, Chang-Shu Wang, and Khalil Sultanem. “Data from Head-Neck-PET-CT.” The Cancer Imaging Archive, 2017. Accessed May 14, 2021. https://wiki.cancerimagingarchive.net/x/24pyAQ.

